# Biallelic variants in *SREK1* downregulating *SNORD115* and *SNORD116* cause a novel Prader-Willi-like syndrome

**DOI:** 10.1101/2025.02.26.24313254

**Authors:** Sadia Saeed, Anna-Maria Siegert, YC. Loraine Tung, Roohia Khanam, Qasim M. Janjua, Jaida Manzoor, Mehdi Derhourhi, Bénédicte Toussaint, Brian Y.H. Lam, Sherine Awad Mahmoud, Emmanuel Vaillant, Emmanuel Buse Falay, Souhila Amanzougarene, Hina Ayesha, Waqas I. Khan, Nosheen Ramazan, Vladimir Saudek, Stephen O’Rahilly, Anthony P. Goldstone, Muhammad Arslan, Amélie Bonnefond, Philippe Froguel, Giles S.H. Yeo

**Author notes:** Corresponding authors: Philippe Froguel, Department of Metabolism, Digestion and Reproduction, Imperial College London, London, UK 0033 3740 08101, Giles S.H. Yeo, Medical Research Council Metabolic Diseases Unit, Institute of Metabolic Science, University of Cambridge, Cambridge, United Kingdom, 0044 1223 769039. These authors contributed equally to the paper and should be considered co-first authors (Sadia Saeed, Anna-Maria Siegert, YC. Loraine Tung). These are joint senior authors (Philippe Froguel, Giles S.H. Yeo).

## Abstract

Up to 10% of patients with severe early-onset obesity carry pathogenic variants in known obesity-related genes, mostly affecting the leptin-melanocortin pathway. Studying children with severe obesity from consanguineous populations provides a unique opportunity to uncover novel molecular mechanisms. Using whole-exome sequencing, followed by a rigorous analytical and filtration strategy, we identified three different homozygous missense variants in *SREK1* (encoding Splicing Regulatory glutamic acid and lysine rich protein) in children with severe obesity, from three unrelated consanguineous pedigrees. The wild type *SREK1* gene of human induced pluripotent stem cell (iPSC)-derived hypothalamic neurons was individually replaced by each of the three variants and the impact of these changes on global gene expression was studied. Neurons expressing the two variants in the *SREK1* RNA recognition domain p.P95L and p.T194M, but not the C-terminally located p.E601K, had markedly reduced expression of the small nucleolar RNA clusters *SNORD115* and *SNORD116*, deficiency of which has been implicated in Prader-Willi syndrome (PWS). In addition to hyperphagic obesity the carriers of these two variants had other features of PWS, such as neonatal hypotonia. In conclusion, homozygous variants in *SREK1* result in a subtype of severe early onset obesity sharing features with PWS.

## INTRODUCTION

Biallelic variants in several genes have been identified to cause severe early-onset obesity. Genetic screening of the Severe Obesity in Pakistani Population (SOPP) cohort, which encompasses children with extreme hyperphagic obesity and their family members from consanguineous pedigrees in Pakistan, have allowed a successful genetic diagnosis in almost 50% of cases (1–8). This predominantly includes pathogenic variants in genes encoding components of the hypothalamic leptin-melanocortin pathway (e.g. *LEP, LEPR* and *MC4R*), crucial regulators of appetite and energy homeostasis (9).

There are also several well-known Mendelian disorders such as Bardet-Biedl, Alström, and Prader-Willi syndromes, which have obesity as one of their hallmark traits accompanying a range of other clinical features, including intellectual disability, dysmorphic features, vision and hearing impairment, maladaptive behaviour, and organ-and cell-specific anomalies(10). Among these, Prader-Willi syndrome (PWS) results from the deletion of a maternally imprinted region on paternal chromosome 15q11-q13 (11), which we have identified in a number of cases within the SOPP cohort (6). We have also previously demonstrated that paternal deletions as small as 187 kb within the PWS region, encompassing the SNORD116 snoRNA cluster, are sufficient to result in PWS, as also seen in other cases with microdeletions in this region (12–15). Beyond known obesity genes, the SOPP cohort has been a valuable resource for the discovery of variants in new genes not previously implicated in weight control including *ADCY3* (8) and *P4HTM* (16).

To uncover new genes associated with obesity, here we have analysed whole exome sequencing (WES) data from patients within SOPP with genetically unexplained obesity. Using the Pakistan Risk of Myocardial Infarction Study (PROMIS) cohort (17), whose participants share a geographically similar background as a control, we assessed the burden of homozygous, rare and potentially deleterious variants in SOPP that significantly increased the risk of obesity. We followed a stepwise filtering strategy, which led to the discovery of homozygous missense variants in *SREK1* (encoding splicing regulatory glutamic acid and lysine-rich protein 1) in unrelated families with severe obesity. These variants were then functionally analysed in human induced pluripotent stem cells (iPSCs) that had been differentiated into hypothalamic neurons. The results showed a significant downregulation of the non-coding RNA clusters, *SNORD115* and the aforementioned *SNORD116,* both of which are located in the maternally imprinted PWS region on chromosome 15q11-13. Assessment of the probands carrying these *SREK1* variants show a significant overlap of phenotypes with that seen in PWS. Thus, we report here that homozygous variants in *SREK1* result in a subtype of severe early onset obesity sharing features of PWS.

## RESULTS

### Comprehensive Evaluation of Severe Obesity in the SOPP Cohort

Using whole exome sequencing data from 463 patients with severe obesity from the SOPP cohort and 1,000 controls from the PROMIS cohort, we detected genes harbouring a burden of rare potentially deleterious variants that significantly increased the risk of obesity using an adjusted MiST analysis (18). Additional filtration criteria included at least three families carrying potentially deleterious variants in the same gene, and enrichment of its expression in the human brain. The candidate variant was further validated through Sanger sequencing of available family members to confirm its segregation with the disease within each family. This comprehensive approach led to the identification of *SREK1* (NM_001077199.3) with significant association between the burden of rare, homozygous, and potentially deleterious variants in this gene, and increased obesity risk (*P*_π_ = 0.021; π^ = 14; *P*_τ_ = 1.0). Three homozygous missense variants in *SREK1*, a c.284C>T variant encoding p.P95L, a c.581C>T variant encoding p.T194M, and a c.1801G>A variant encoding p.E601K, were identified in three unrelated pedigrees (Figure 1).

**Figure 1:**
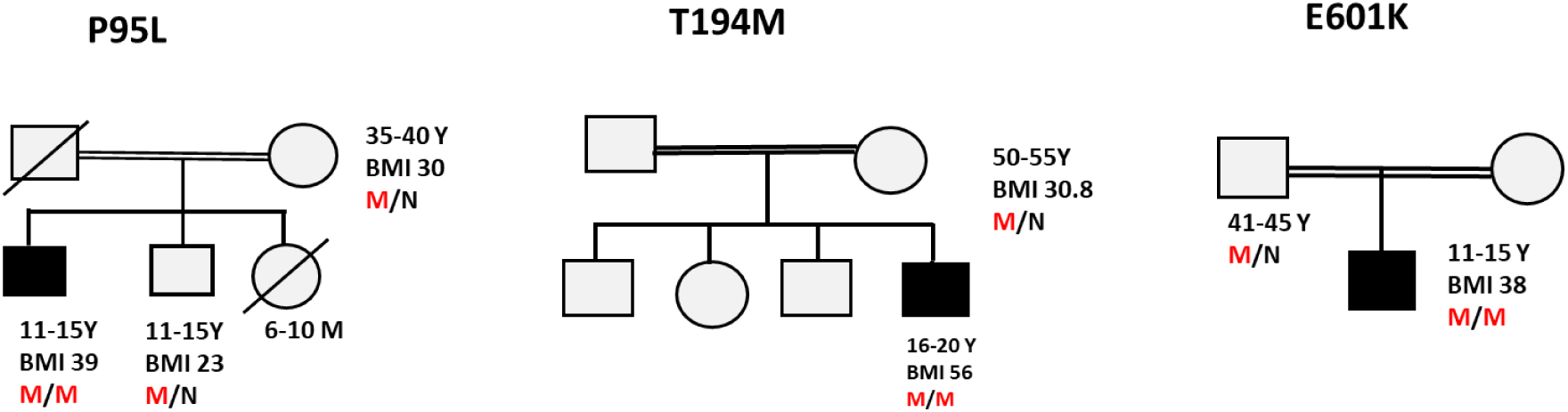
Pedigrees of families with *SREK1* mutations. Family trees of three consanguineous pedigrees with individuals carrying *SREK1* variants (P95L, T194M, and E601K).

### Variant identification and analysis

*SREK1* exhibits extremely high genetic constraint, particularly with a probability of loss-of-function (pLoF) score of 1.00 as reported in GnomAD v4.1.0, indicating a critical intolerance to high-impact functional variants. The observed single nucleotide variants (SNVs) are also significantly lower than expected, with 11 observed, compared to 59.7 expected, underscoring that functional disruption of this gene is likely to have severe consequences.

The c.284C>T variant, encoding p.P95L, is a novel variant that has not been reported in any existing databases, including ClinVar, dbSNP, the 1000 Genomes Project, and as reported in GnomAD v4.1.0. Computational predictive analyses indicate its potential pathogenicity, with a PolyPhen score of 0.997 (probably damaging) and SIFT score of 0.00 (deleterious).

The c.581C>T variant, encoding p.T194M, has a minor allele frequency of 1.3 × 10^-4^ in South Asians (gnomAD v4.1.0) but has never been reported in the homozygous state. Computational predictive analyses suggest its potential pathogenicity, with a PolyPhen score of 0.997 (probably damaging) and SIFT score of 0.03 (deleterious).

The c.1801G>A variant encoding p.E601K, also has a minor allele frequency of 1.3 × 10^-4^ in South Asians (gnomAD v4.1.0) but has never been reported in the homozygous state. It was assessed with a PolyPhen score of 0.969 (probably damaging) and a SIFT score of 0.00 (deleterious).

### Clinical history of patients with *SREK1* variants

All individuals carrying a homozygous variant in *SREK1* exhibited hyperphagic obesity and neurodevelopmental delay. Conversely, all available family members who were heterozygous for these variants did not display early-onset severe obesity, neurodevelopmental delay or related abnormalities (Figure 1).

### Case 1 - variant encoding p.P95L

The patient, carrying the homozygous variant encoding p.P95L, was a male initially recruited at 6-10 years of age, presenting with obesity (body mass index [BMI] 29 kg/m²). Additional symptoms included hyperphagia, aggressive behaviour and below-average academic performance. By age 11-15 years, his BMI had increased to 39 kg/m² with a weight of 93 kg. At 16-20 years, his BMI further rose to 48 kg/m², and his weight reached 124 kg.

### Case 2 - variant encoding p.T194M

The patient, carrying the homozygous variant encoding p.T194M, was a male initially recruited at 16- 20, presenting with a BMI 56 kg/m² and weight 169 kg. At the time of presentation, the patient exhibited psychological and behavioural challenges in addition to obesity. Despite these challenges, he benefited from psychological support and counselling. With the aid of a strict diet and gym training, by age 26-30 years, he reduced his BMI to 42 kg/m².

### Case 3 - variant encoding p.E601K

The patient, carrying the homozygous variant encoding p.E601K was a male first seen when he was 11-15 years old, presenting with BMI 38 kg/m² and weight 80 kg. Clinical features included social withdrawal, intellectual performance about 2 years behind chronological age, and social anxiety. At the three-year follow-up from his initial recruitment, his BMI was 41 kg/m², and his weight was 118 kg.

### Heterozygous *SREK1* null variants not associated with BMI in UK Biobank

Using exome sequencing data from up to 187,242 participants in the UK Biobank, we investigated the association between null variants in *SREK1* (Supplemental Table 1) and obesity-related traits using adjusted MiST analyses. We found no significant association between these null variants (n=5) and obesity-related traits. Specifically, for higher BMI levels, the results were as follows: Pπ = 0.89; π^ = 0.01; Pτ = 0.75. For overweight levels, the findings were: Pπ = 0.51; π_hat = 0.72; Pτ = 0.21. It is, however, important to note that all detected null variants in the UK Biobank were heterozygous.

### Structural analysis of SREK1 domains

SREK1 belongs to a family of serine/arginine rich splicing factors (SRFS) as its 13^th^ member. AlphaFold and InterPro databases reveal that it consists of two well defined RNA recognition motifs (RRM), followed by multiple intermittent regions containing serine/arginine (SR) rich and glutamic (or aspartic) acid/lysine-rich sequences (EK) sequences. The arrangement of these components within the primary protein structure is depicted in Figure 2. They are known to be responsible for interaction with specific nucleotide sequences within the RNA molecules. SREK1 is present in the genomes of all metazoans and its RRM domains are extremely well conserved, suggesting that they bind to very specific RNA sequences. Two of the variants, encoding p.P95L and p.T194M, are situated within the RRM domains (Figure 2). The EK segment folds into long α-helices with regular distribution of positive and negative charges, suggesting that they probably interact non-specifically with RNA double helix stems. The SR segments, in which many serines are phosphorylatad (phosphoSitePlus database), are regions of low complexity, characterized by biased amino acid compositions, and containing short repetitive sections enriched in one or several amino acids. They often act as scaffolding domains, usually disordered on their own, but structured in interaction with other proteins. They nucleate and control formation of larger macromolecular complexes, they can also interact with other proteins at its opposite interface, as demonstrated when it is embedded within a larger protein/RNA complex (19). The variant encoding p.E601K variant is situated outside of these well-understood functional domains.

**Figure 2:**
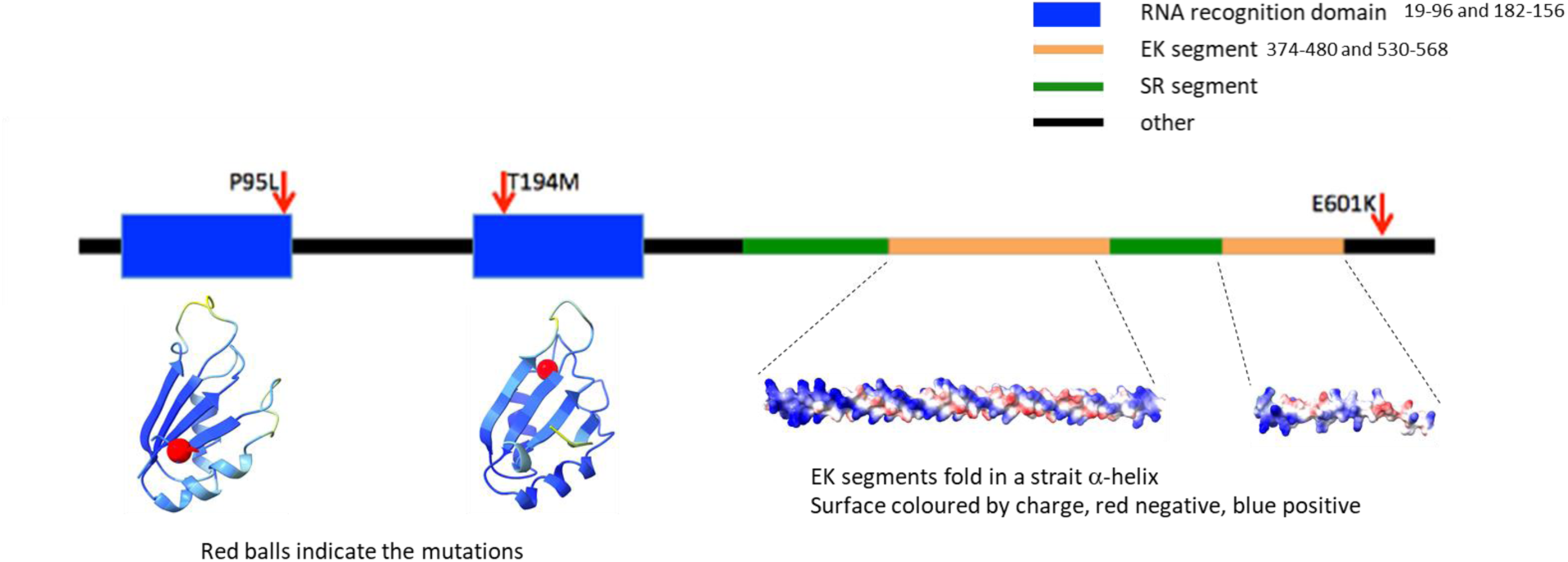
**Domain structure and variants of *SREK1*** A schematic illustration with the AlphaFold-predicted structure of the SREK1 domains, highlighting the distinct feature of the EK segments folding into a straight α-helix with regular surface charge distribution. The RNA recognition motifs (residues 19–96 and 173–256) are color-coded based on AlphaFold’s quality prediction convention (deep blue for very high confidence, light blue for high confidence).

### Gene expression analysis of *SREK1* mutants in iPSC-derived hypothalamic neurons

Genetic studies point to the brain, and in particular the hypothalamus, as having a crucial leading role in modulating appetitive behaviour (20), so we chose to functionally characterise these *SREK1* variant in a human neuronal context. Human induced pluripotent stem cell (iPSC)-derived hypothalamic neurons recapitulate many essential properties of *in vivo* counterparts, thus making them a relevant human model to study obesity-related genetic variants, as we have previously reported (20). We introduced the *SREK1* variants into human iPSCs using CRISPR/Cas9 gene editing and then differentiated them into neurons as previously described (21). We then performed bulk RNA sequencing on these iPSC-differentiated hypothalamic neurons.

Multidimensional scaling (MDS) analysis showed the two variants encoding p.P95L and p.T194M, that are situated within the RRM domains, showed very similar transcriptomic fingerprints, as compared to the WT and E601K neurons (Figure 3A). When we performed differential gene expression analyses to identify all differentially expressed genes across the four genotypes (n=1315, FDR p<0.05), and performed unsupervised hierarchical clustering, the p.P95L and p.T194M expressing neurons unequivocally cluster together, distinct from the WT and p.E601K neurons, which themselves cluster together (Figure 3B). When we grouped the p.P95L and p.T194M expressing neurons together and compared these to WT neurons, amongst the differentially expressed transcripts were multiple small nucleolar RNAs including those belonging to the *SNORD115* and *SNORD116* families (Figure 3C). While there was a similar pattern when we compared the p.E601K expressing neurons with WT, none of these reached statistical significance (Figure 3D, FDR <0.05).

**Figure 3:**
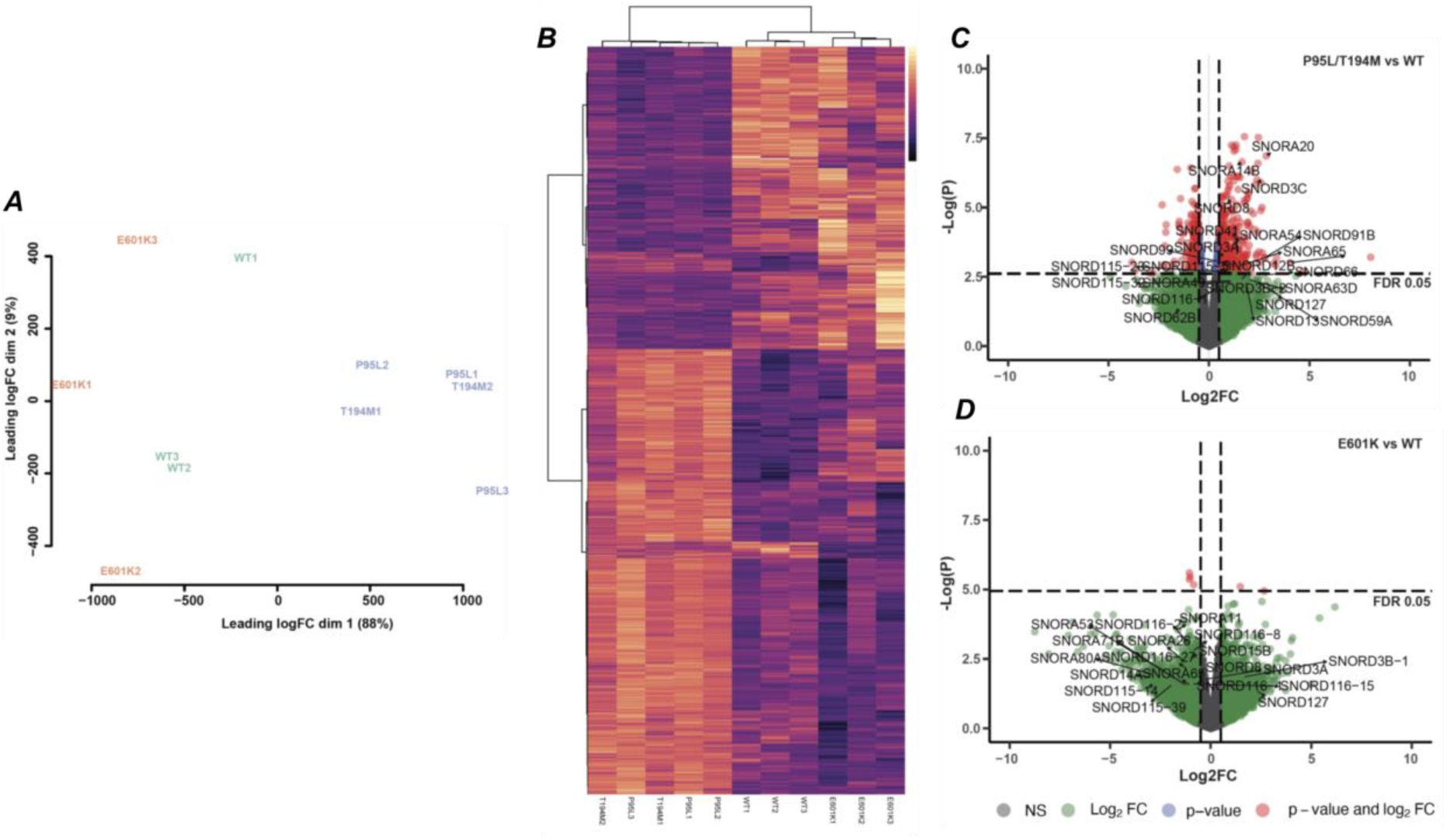
RNAseq results for *SREK1* RNA recognition domain and p.E601K variants. **(A)** Principal Component Analysis (PCA) plots of RNAseq data comparing wild-type (WT) *SREK1* and variants. The clustering of dots between the RNA recognition domain variants indicates the similarity between these samples. **(B)** Heatmap of differentially expressed genes (DEGs) that visualises the changes in gene expression associated with different variants in the *SREK1* gene. The colour intensity corresponds to the level of gene expression, with darker colours indicating stronger expression changes. **(C) & (D)** Volcano plots indicating DEGs between the RNA recognition domain variants (p.P95L and p.T194M) vs WT and p.E601K vs WT control, respectively. The Differential expression analysis of the RNA recognition domain variants compared to the wild type highlighted the dysregulation of several small nucleolar RNAs (snoRNAs), particularly in the *SNORD115* and SNORD116 families.

Because all of these small nucleolar RNAs are only approximately 100 nucleotides in length, and our RNAseq approach was not optimised for such short transcripts, we designed a qRT-PCR assay to specifically assess the expression of *SNORD115* and *SNORD116*. We show that *SNORD115* and *SNORD116* are downregulated in neurons expressing the RRM domain variants, but not in those with the C-terminal variant encoding p.E601K (Figure 4). We previously reported that deletions encompassing the *SNORD116* region on chromosome 15q11 implicate this cluster of non-coding RNAs in a causative role for PWS (15), as also reported by others (12–14). Additionally, we have previously reported that hypothalamic specific deletion of *Snord116* in adult mice recapitulates the hyperphagia of PWS (22).

**Figure 4:**
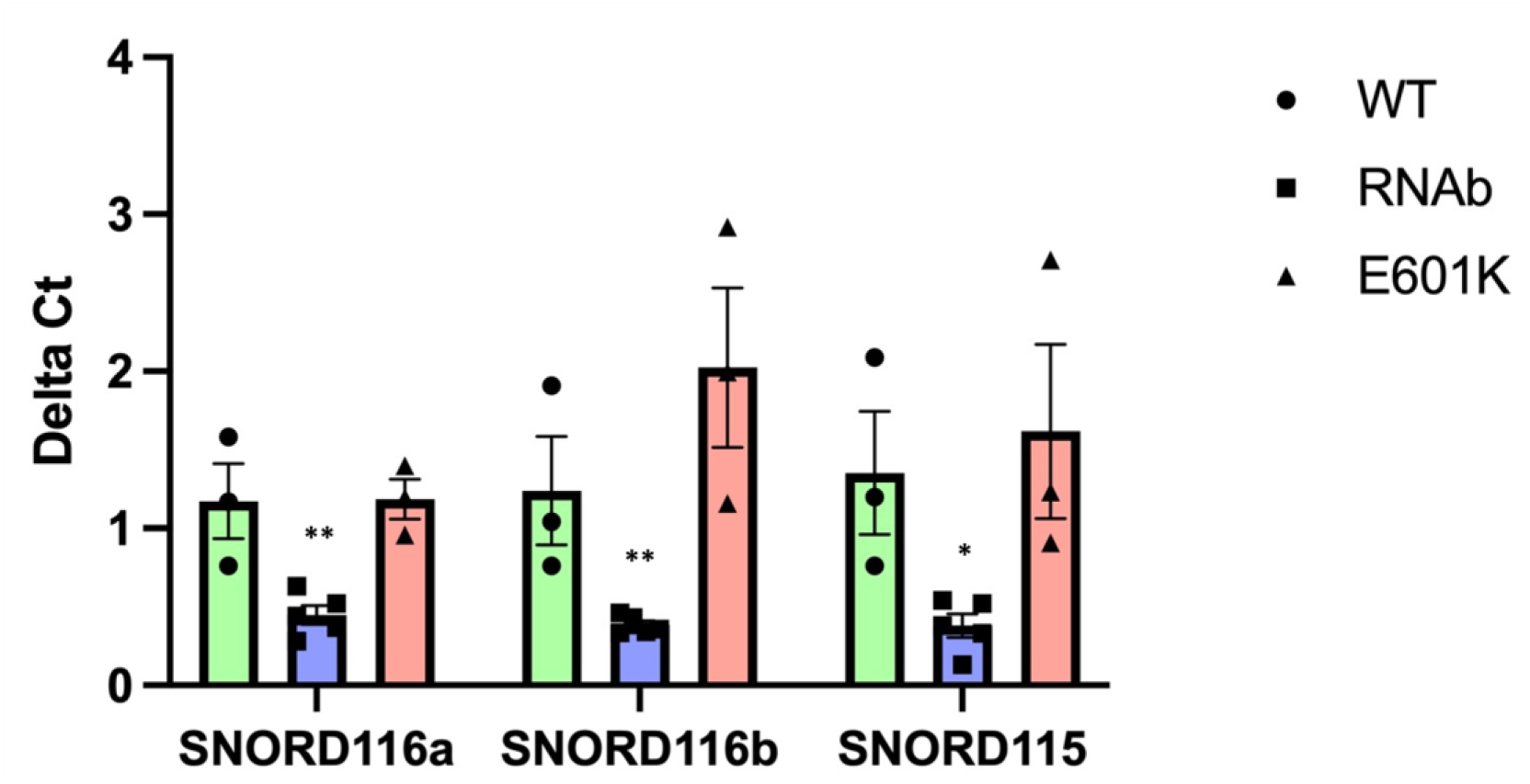
RT-PCR verification of *SNORD116* and *SNORD115* changes in *SREK1* RNA recognition domain and p.E601K variant. The mRNA levels of both group I (*SNORD116a*), group II (*SNORD116b*) *SNORD116* clusters and *SNORD115* were significantly reduced in the RNA recognition domain variant samples compared to the wild-type (WT) control. The expression levels were normalised to *GAPDH*. Data are presented as mean ± SEM and statistical significance was calculated using multiple unpaired *t*-test based on data from at least three independent experiments (**p<0.01, *p<0.05).

Thus, our results suggest that variants in the RNA recognition domain of *SREK1* lead to a downregulation of *SNORD115* and *SNORD116* that could play a role in the phenotype of severe obesity seen in the probands.

### Evaluation of phenotypic overlap with PWS

Given the finding that variants in the RNA recognition motif of SREK1 resulted in the downregulation of *SNORD115* and *SNORD116*, both implicated in the hyperphagia and weight gain characteristics of PWS, we proceeded to assess the potential overlap of phenotypes between carriers with *SREK1* RRM variants and PWS.

Unfortunately, consent for the follow-up detailed comparison using the PWS questionnaire and cognitive assessment was not obtainable from the parent of the proband carrying the p.E601K variant, preventing such a comparison. However, in June 2023, updated anthropometric measurements and data from the Dykens hyperphagia questionnaire were collected during the hospital visits for the two probands carrying the variants encoding p.P95L or p.T194M. Both probands demonstrated moderate hyperphagia from childhood, with their scores on the Dykens hyperphagia questionnaire falling within the upper half of the range typically observed in children and adults with PWS (23, 24). Specifically, their total scores ranged from 37 to 39 (with a possible minimum of 11 and a maximum of 55), and their scores on the sub-scales of hyperphagic behaviour, drive and severity were similarly elevated (Table 1, 2).

**Table 1:**
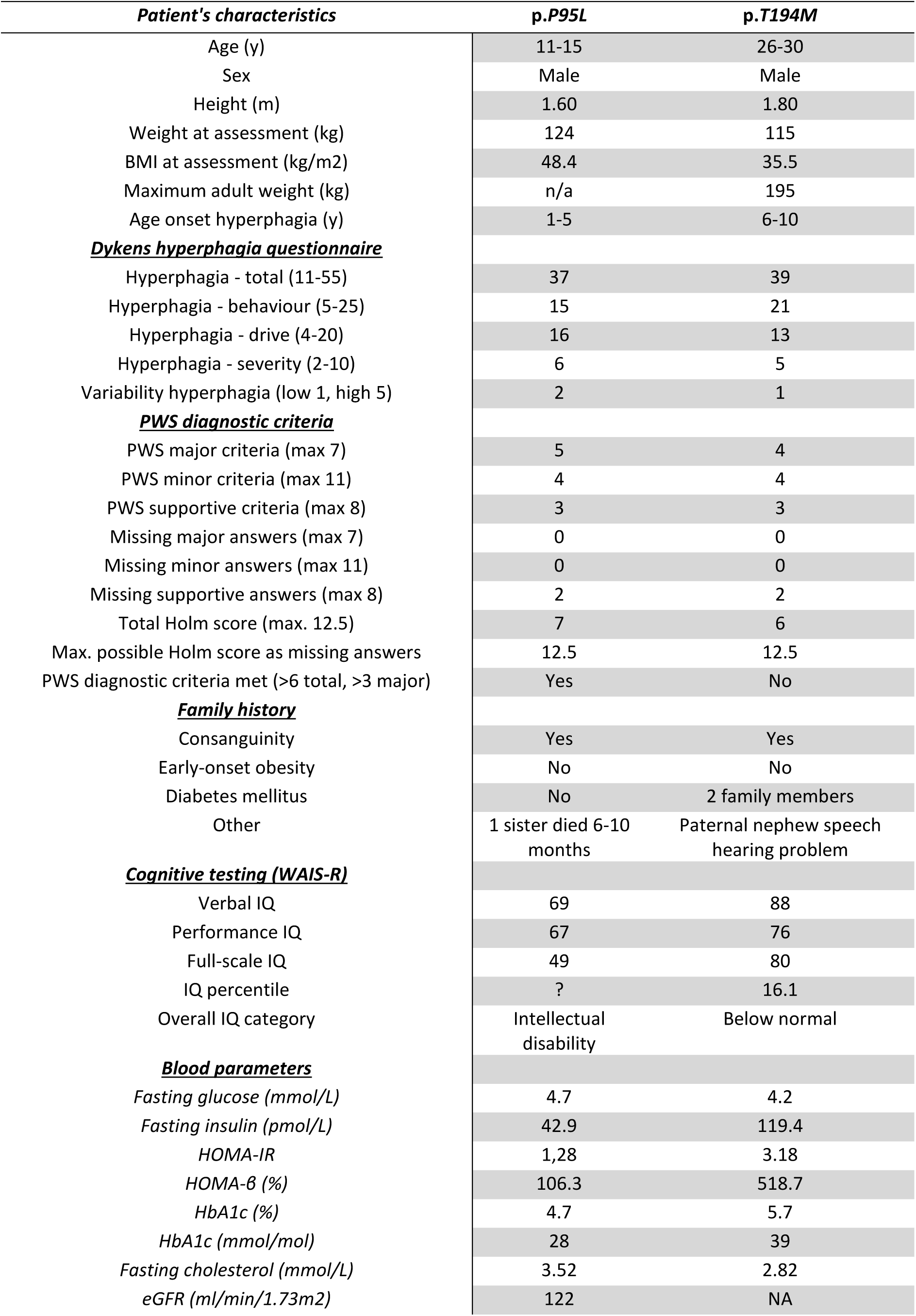
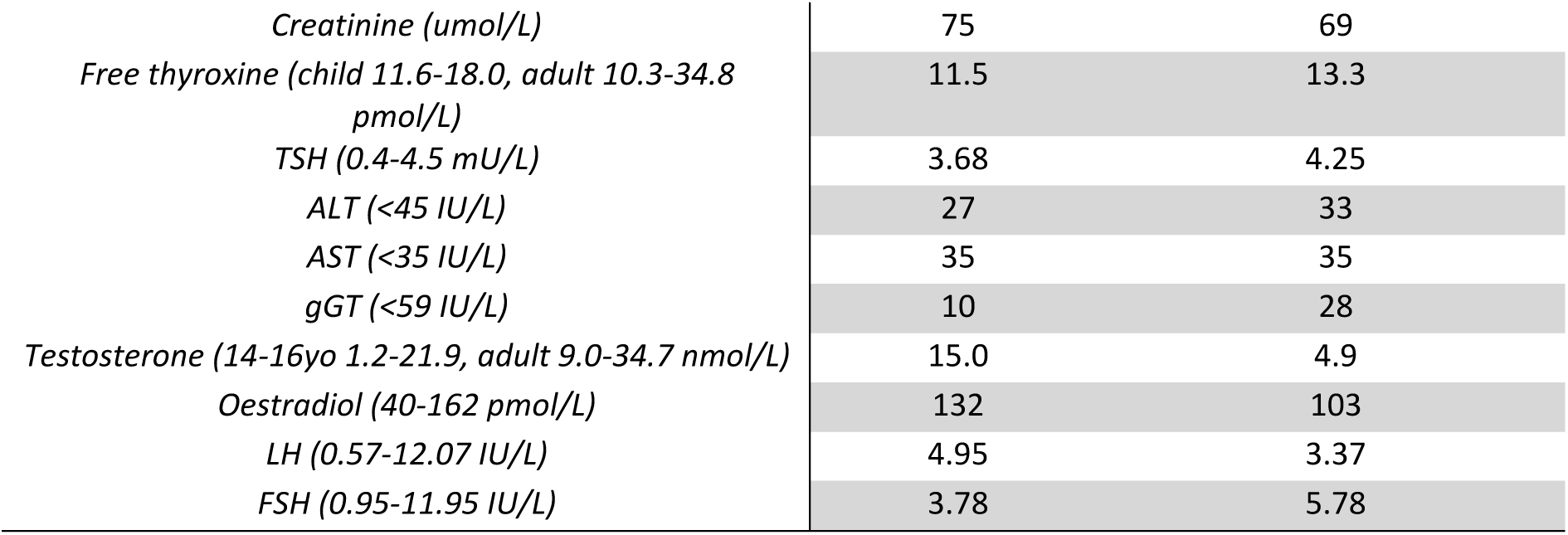
Clinical, familial and cognitive characteristics of patients with biallelic *SREK1* variants (p.P95L and p.T194M)

| *Patient's characteristics* | *p.P95L* | *p.T194M* |
| --- | --- | --- |
| Age (y) | 11-15 | 26-30 |
| Sex | Male | Male |
| Height (m) | 1.60 | 1.80 |
| Weight at assessment (kg) | 124 | 115 |
| BMI at assessment (kg/m<sup>2</sup>) | 48.4 | 35.5 |
| Maximum adult weight (kg) | n/a | 195 |
| Age onset hyperphagia (y) | 1-5 | 6-10 |
| **<u>Dykens hyperphagia questionnaire</u>** |  |  |
| Hyperphagia - total (11-55) | 37 | 39 |
| Hyperphagia - behaviour (5-25) | 15 | 21 |
| Hyperphagia - drive (4-20) | 16 | 13 |
| Hyperphagia - severity (2-10) | 6 | 5 |
| Variability hyperphagia (low 1, high 5) | 2 | 1 |
| **<u>PWS diagnostic criteria</u>** |  |  |
| PWS major criteria (max 7) | 5 | 4 |
| PWS minor criteria (max 11) | 4 | 4 |
| PWS supportive criteria (max 8) | 3 | 3 |
| Missing major answers (max 7) | 0 | 0 |
| Missing minor answers (max 11) | 0 | 0 |
| Missing supportive answers (max 8) | 2 | 2 |
| Total Holm score (max. 12.5) | 7 | 6 |
| Max. possible Holm score as missing answers | 12.5 | 12.5 |
| PWS diagnostic criteria met (>6 total, >3 major) | Yes | No |
| **<u>Family history</u>** |  |  |
| Consanguinity | Yes | Yes |
| Early-onset obesity | No | No |
| Diabetes mellitus | No | 2 family members |
| Other | 1 sister died 6-10 months | Paternal nephew speech hearing problem |
| **<u>Cognitive testing (WAIS-R)</u>** |  |  |
| Verbal IQ | 69 | 88 |
| Performance IQ | 67 | 76 |
| Full-scale IQ | 49 | 80 |
| IQ percentile | ? | 16.1 |
| Overall IQ category | Intellectual disability | Below normal |
| **<u>Blood parameters</u>** |  |  |
| Fasting glucose (mmol/L) | 4.7 | 4.2 |
| Fasting insulin (pmol/L) | 42.9 | 119.4 |
| HOMA-IR | 1,28 | 3.18 |
| HOMA-β (%) | 106.3 | 518.7 |
| HbA1c (%) | 4.7 | 5.7 |
| HbA1c (mmol/mol) | 28 | 39 |
| Fasting cholesterol (mmol/L) | 3.52 | 2.82 |
| eGFR (ml/min/1.73m<sup>2</sup>) | 122 | NA |
| *Creatinine (umol/L)* | 75 | 69 |
| *Free thyroxine (child 11.6-18.0, adult 10.3-34.8 pmol/L)* | 11.5 | 13.3 |
| *TSH (0.4-4.5 mU/L)* | 3.68 | 4.25 |
| *ALT (<45 IU/L)* | 27 | 33 |
| *AST (<35 IU/L)* | 35 | 35 |
| *gGT (<59 IU/L)* | 10 | 28 |
| *Testosterone (14-16yo 1.2-21.9, adult 9.0-34.7 nmol/L)* | 15.0 | 4.9 |
| *Oestradiol (40-162 pmol/L)* | 132 | 103 |
| *LH (0.57-12.07 IU/L)* | 4.95 | 3.37 |
| *FSH (0.95-11.95 IU/L)* | 3.78 | 5.78 |

**Table 2:** Comparative analysis of hyperphagia across biallelic *SREK1*, biallelic *LEPR* and heterozygous *MC4R* variants, and PWS, using Dykens Hyperphagia Questionnaire.

| Dyken's Hyperphagia Questionnaire | *SREK1*<br>p.P95L | *SREK1*<br>p.T194M | *LEPR*<br>(n=8) | *MC4R*<br>(n=7) | PWS<br>(n=21) | PWS<br>(n=24) | PWS<br>(n=21) | PWS<br>(n=100) |
| --- | --- | --- | --- | --- | --- | --- | --- | --- |
| Reference no. |  |  | (24) | (24) | (47) | (48) | (49) | (50) |
| Age (years) | 11-15 | 26-30 | 7.1 ± 5.7 | 10.7 ± 6.2 | 15.5 ± 6.6 | 26.3 ± 6.9 | 27.8 ± 8.3 | 28.2 ± 7.7 |
| Sex | Male | Male | Male & female | Male & female | Male & female | Male & female | Male & female | Male & female |
| BMI at assessment (kg/m<sup>2</sup>) | 48.4 | 35.5 | 45.3 ± 15.9 | 35.1 ± 10.0 | 24.3 ± 6.8 | 30.2 ± 6.4 | 30.5 ± 6.3 | 42.0 ± 10.6 |
| Hyperphagia - behaviour (5-25) | 15 | 21 | 13.4 ± 3.9 | 11.9 ± 2.3 | 16.4 ± 3.9 | 14.1 ± 5.8 | 14.1 ± 5.2 | 12.8 ± 4.3 |
| Hyperphagia - drive (4-20) | 16 | 13 | 13.1 ± 3.6 | 14.0 ± 0.8 | 13.6 ± 3.7 | 11.2 ± 4.2 | 10.9 ± 3.7 | 11.5 ± 3.3 |
| Hyperphagia - severity (2-10) | 6 | 5 | 5.5 ± 2.9 | 5.1 ± 2.3 | 7.7 ± 1.5 | 4.5 ± 2.6 | 4.6 ± 2.7 | 4.6 ± 2.1 |
| Hyperphagia - total (11-55) | 37 | 39 | 32.0 ± 9.3 | 21.4 ± 5.5 | 39.7 ± 11.8 | 29.8 ± 10.1 | 29.6 ± 9.5 | 28.9 ± 8.0 |

Data given as mean ± SD. Data for patients with biallelic *LEPR* and heterozygous *MC4R* variants taken from (24) and for PWS taken from (47-50)

Only the proband carrying the variant encoding p.P95L, now 11-15-years old, met all the major clinical diagnostic criteria for PWS (Table 3) (25). The proband with the variant encoding p.T194M, now 26-30-years old, lacked one major criterion necessary for a definitive clinical diagnosis as the onset of obesity occurred after surpassing the diagnostic cut-off age of 6 years (25).

**Table 3:** Clinical features of major PWS diagnostic criteria in patients with biallelic *SREK1* variants (p.P95L and p.T194M)

| Major PWS Diagnostic Criteria | p.P95L | p.T194M |
| --- | --- | --- |
| Neonatal hypotonia | Yes | Yes |
| Poor suck | Yes | Yes |
| Challenge feeding at birth | Yes | Yes |
| Poor weight gain in infancy | No | Yes |
| Obesity | Yes | Yes |
| Age onset (y) | 1-5 | 6-10 |
| Hyperphagia | Yes | Yes |
| *Facial appearance* |  |  |
| Increased skull AP dimension | No | No |
| Narrow bifrontal diameter | No | Yes |
| Small mouth with thin upper lip | Yes | Yes |
| Down-turned corners mouth | Yes | No |
| Almond shaped palpebral fissure | No | No |
| Hypogonadism | No | No |
| Hypogenitalism | No | No |
| Learning disability | Yes | No |
| Global developmental delay <6y | Yes | Yes |

Both probands also exhibited some characteristic facial features, such as small mouth with a thin upper lip, with one subject each showing a narrow bifrontal diameter and downturned corners of the mouth (Table 4). Only the proband with the variant encoding p.P95L met criteria for intellectual disability (WAIS-R full-scale intelligence quotient (IQ) 49), but the proband with the variant encoding p.T194M was of low normal intelligence (full-scale IQ 80, 16^th^ percentile). Both probands had a history of behavioural problems characteristic for PWS, including temper tantrums, violent outbursts, obsessive / compulsive behaviours. Additionally, both probands reported a history of anxiety, particularly around access to food, and the 26-30 years old proband carrying the variant encoding p.T194M had a history of depression, but there was no history of psychosis (Table 5).

**Table 4:** Clinical features of minor PWS diagnostic criteria in patients with biallelic *SREK1* variants (p.P95L and p.T194M)

| Minor PWS Diagnostic Criteria | p.P95L | p.T194M |
| --- | --- | --- |
| Lethargy as infant | Yes | Yes |
| Temper tantrums | Yes | Yes |
| Violent outbursts | Yes | Yes |
| Obsessive / compulsive behaviours | Yes | Yes |
| Tendency to be argumentative, oppositional, rigid, manipulative, possessive, stubborn | Yes | Yes |
| Perseverates | Yes | Yes |
| Steals | NA | Yes |
| Lies | Yes | Yes |
| Disturbed or noisy sleep | Yes | No |
| Sleep apnoea | Yes | Yes |
| *Physical appearance* |  |  |
| Short stature by 15y | No | No |
| Fair skin or hair | Yes | Yes |
| Small hands | No | No |
| Small feet | No | No |
| Narrow hands with straight ulnar border | No | No |
| *Eye related issues* |  |  |
| Myopia | No | No |
| Esotropia (squint) | No | No |
| Thick saliva with crusting at mouth corners | No | No |
| Speech articulation defects | No | No |
| Skin picking | No | No |

**Table 5:**
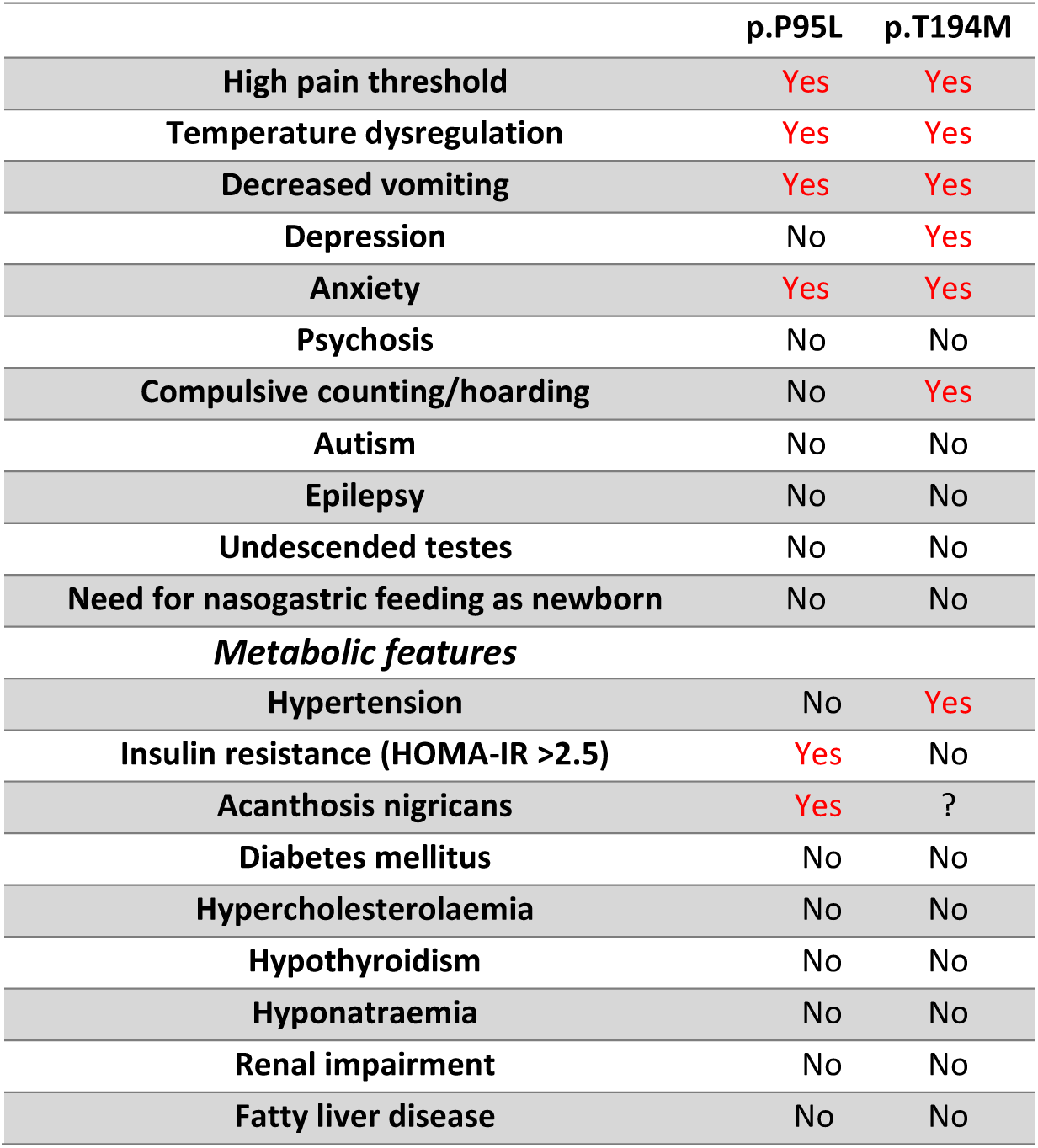
Clinical features of supportive PWS diagnostic criteria and other neuropsychological and metabolic phenotypes in patients with biallelic *SREK1* variants (p.P95L and p.T194M)

|  | p.P95L | p.T194M |
| --- | --- | --- |
| High pain threshold | Yes | Yes |
| Temperature dysregulation | Yes | Yes |
| Decreased vomiting | Yes | Yes |
| Depression | No | Yes |
| Anxiety | Yes | Yes |
| Psychosis | No | No |
| Compulsive counting/hoarding | No | Yes |
| Autism | No | No |
| Epilepsy | No | No |
| Undescended testes | No | No |
| Need for nasogastric feeding as newborn | No | No |
| *Metabolic features* |  |  |
| Hypertension | No | Yes |
| Insulin resistance (HOMA-IR >2.5) | Yes | No |
| Acanthosis nigricans | Yes | ? |
| Diabetes mellitus | No | No |
| Hypercholesterolaemia | No | No |
| Hypothyroidism | No | No |
| Hyponatraemia | No | No |
| Renal impairment | No | No |
| Fatty liver disease | No | No |

Neither proband exhibited short stature nor small hands or feet. Stimulation tests for presence of growth hormone deficiency or measurement of serum IGF-1 were unavailable for either proband. Both probands reported a high pain threshold and temperature dysregulation, though there was no history of skin picking, speech articulation defects or strabismus. Both probands reported decreased vomiting, but neither experienced any other gastrointestinal or genitourinary problems, such as constipation or bladder instability, that are often observed in PWS.

Both probands had a history of sleep apnoea related to obesity. The proband with the variant encoding p.P95L exhibited insulin resistance, though he did not develop type 2 diabetes mellitus. The proband with the variant encoding p.T194M only experienced hypertension. He effectively lowered his BMI from 60.2 kg/m² to 35.5 kg/m² over a period of 7 years by following dietary recommendations from a dietitian.

## DISCUSSION

In this study, we identified a novel form of childhood-onset monogenic severe obesity caused by homozygous missense variants in *SREK1*, a gene not previously associated with body weight regulation. Although the clinical features of individuals carrying a single homozygous variant in *SREK1* surprisingly resemble certain aspects of PWS, which is most commonly caused by deletions on the paternal chromosome 15q11-q13, or chromosome 15 maternal uniparental disomy, our functional study using human iPSC-derived hypothalamic neurons provided further insight. This study demonstrated downregulation of *SNORD115* and *SNORD116* expression in iPSC-derived hypothalamic neurons in two of the three cases of homozygous *SREK1* missense variants, establishing a molecular link that explains the occurrence of PWS-like obesity from independent families.

Clinically, the relatively late age-of-onset of obesity and hyperphagia in both probands mirrors findings in PWS (25). This stands in stark contrast to what is seen with loss-of-function rare biallelic variants in leptin (*LEP*) and leptin receptor (*LEPR*), where obesity initiates before 2 years of age (26–28). Hypotonia is present in both *SREK1* variant carriers and individuals with PWS, though the phenotypic overlap is not absolute with the p.T194M carrier proband not meeting all the classic clinical criteria for a PWS diagnosis. Notably, 17% of patients with a molecular diagnosis of PWS did not fulfil the complete clinical diagnostic criteria (29). Many features of PWS are indeed subtle or non-specific and evolve with age. However, based on the Dykens Hyperphagia Questionnaire (23), hyperphagia is evident in our probands with variants within the RRM domain. Their scores are comparable or above the mean for those previously reported for patients with PWS in similar age groups (23, 24) (Table 2). In addition, our probands exhibit all the classical manifestation of food-related behavioural problems commonly seen in individuals with PWS, including excessive appetite, overeating, food obsession and a lack of satiation. The total hyperphagia scores for both *SREK1* probands were also higher than the mean score for patients with biallelic variants in *LEPR* or heterozygous variants in *MC4R* (Table 2), as well as that previously reported in other genetic obesity syndromes including WAGR syndrome, Alstrom syndrome, pseudohypoparathyroidism 1A and Bardet-Biedel syndrome (24). This shows the severity of the appetite dysregulation due to biallelic variants in *SREK1*.

Other developmental clinical hallmarks of PWS including neonatal/infantile lethargy and poor suck, as well as delayed physical and social developmental milestones, were also reported in our probands. In contrast, endocrine abnormalities typically seen in PWS were not observed in the *SREK1* probands. Rodent and human studies have suggested loss of paternal *MKRN3* and *MAGEL2* genes within the PWS critical region may contribute to the incomplete and/or delayed pubertal development and premature adrenarche commonly seen in PWS (30, 31), though hypogonadism, GH deficiency and short stature have been reported in patients with paternal *SNORD116* microdeletions (12–15) (Supplemental Figure 1).

The role of *SREK1* variants in obesity and in other associated neurological features is also supported by other strong biological data. Functional characterisation of these variants in human iPSC-derived hypothalamic neurons revealed a dysregulation in the expression of multiple non-coding RNAs. Given the role that SREK1 plays in the spliceosome, it may be not surprising that many of the most significantly altered genes are noncoding RNAs, including snoRNAs, crucial for alternative splicing of pre-mRNA molecules. Beyond splicing, these serine/arginine-rich splicing factors are known to play roles in transcription, export, translation, and RNA decay (32), as evidenced by the enrichment of tRNAs, yRNAs and transcription factors on the most significantly regulated gene list. In particular, multiple families of small nucleolar RNAs (snoRNAs) are downregulated.

Upon deeper investigation we demonstrate that variants in the RNA recognition motifs of SREK1 result in a coordinate downregulation of the small nucleolar RNA clusters *SNORD115* and *SNORD116* (Figure 4), a situation in common with the complex genetic imprinting disorder PWS. Thus, our data in SREK1 deficiency confirm that the downregulation of *SNORD115* and *SNORD116* underlies the characteristic symptoms of severe early onset obesity and hyperphagia, hence linking *SREK1* to a novel condition of PWS-like syndromic obesity.

A CRISPR-generated *Srek1* knockout mutant has been developed by the Knockout Mouse Phenotyping Program (KOMP2) at The Jackson Laboratory, Bar Harbor, ME. The Viability Primary Screen phenotypic assay revealed that homozygous knockouts exhibit preweaning lethality with incomplete penetrance, making body weight assessments challenging. In contrast, heterozygous *Srek1* knockout mice showed no association with body weight phenotype, consistent with our observations from the human UK Biobank data. However, there is frequent discordance between human and rodent data for monogenic development disorders.

While multiple mouse models for *Snord116* and *Snord115* have been studied, particularly in the context of PWS, they do not fully replicate all aspects of the human condition. This highlights further the challenges of using rodent models to study syndromic obesity. Indeed, several patient reports demonstrate that loss of *SNORD116* cluster expression results in a phenotype that substantially overlaps with PWS, including all most of clinical features of PWS (12–15). However, paternal deletion of the *Snord116* cluster in mice exhibited several characteristics of PWS, but they do not display all the neurodevelopmental and endocrine abnormalities seen in human PWS (33–36). Furthermore, studying the contribution of *SNORD116* to metabolic syndrome is challenging because paternal *Snord116* knockout mice display early growth retardation reminiscent of the failure to thrive as seen in humans with PWS. However, unlike in human PWS, these mice do not undergo the phenotypic transition to hyperphagia and obesity as they age. Instead, they remain smaller than their wild-type littermates and do not develop obesity, nor do they show significant increases in food intake.

It does remain to be seen if the E601K *SREK1* variant is causative of the severe obesity. The gene expression analysis of human iPSC derived neurons argues against it, as the down regulation of SNORD115/116 seen in the RRM variants are not seen with this variant. However, due to the lack of phenotypic information from the E601K variant carrier, we are not in a position to make any conclusive statements.

In conclusion, our identification of variants in the *SREK1* gene uncovers a new genetic driver of syndromic form of severe obesity. These variants disrupt the RNA-recognition motif of SREK1, leading to a significant and consistent downregulation of *SNORD115* and *SNORD116*, essential non-coding RNAs implicated in PWS. This novel insight highlights the crucial role of splicing regulation in obesity and uncovers an uncharted genetic pathway contributing to syndromic obesity. Our findings further open the door to innovative therapeutic strategies targeting genes involved in splicing mechanisms, heralding a new era in the fight against severe obesity. Finally, this study further illustrates the power of the genetics of obesity in consanguineous families for the discovery of new pathways involved in weight regulation.

## METHODS

### Consent statement

Informed consent was obtained from the legal representatives of all study participants. This consent covers both participation in the study and the publication of its findings.

### Study population and genetic analysis

All probands with severe obesity from the SOPP cohort (*N*=550) were systematically screened for variants in *LEP* and *MC4R* genes through Sanger sequencing. The pathogenicity of these variants was evaluated in accordance with the guidelines and standards established by the American College of Medical Genetics and Genomics (ACMG) (37). Probands who tested negative for (likely) pathogenic variants in these two genes underwent further analysis through whole-exome sequencing, as detailed in previous studies (6). All genes known to be associated with monogenic (syndromic/non-syndromic) obesity (6) were examined for (likely) pathogenic variants. In cases where we could not identify a potentially causative variant (n=463) in known monogenic (syndromic or non-syndromic) obesity genes, we utilized MiST analysis to uncover novel causative genes.

The MiST method was first published in 2013 (18) and since has been used in several studies that demonstrated that MiST performs best with regard to its statistical power across a range of architectures. Thus, for the next gene-centric analysis, we used the MiST method method (38) to identify a burden of variants significantly increasing the risk of obesity. This analysis included 463 patients with genetically unexplained obesity from the SOPP cohort and 1,000 controls from the Pakistani PROMIS (Pakistan Risk of Myocardial Infarction Study) population (17). Details of the analysis are provided elsewhere (16). This approach enables the identification of genes that carry a significant burden of rare (i.e., minor allele frequency below 1%), homozygous, and potentially deleterious variants (as determined by SIFT and PolyPhen) among the obesity cases from the SOPP. Following the MiST analysis, additional filtration criteria were applied, including identification in three or more families, segregation of the variant with the disease within the family, and predominant expression of candidate genes in the human brain based on GTEx (39) and Hypomap data (40).

### MiST analysis on *SREK1* variants

Rare variants in the *SREK1* gene were identified using whole exome sequencing data from the UK Biobank, including 187,242 exome-sequenced samples. This study was conducted under UK Biobank application #67575, approved by the North West Multi-centre Research Ethics Committee (MREC). Variants with a MAF below 1% were included, with a focus on null variants defined as nonsense, frameshift, or canonical splice site variants. Only variants with a read depth >10 and a genotype quality (GQ) score >20 were considered for further analysis. The association between *SREK1* null variants and obesity was assessed using the MiST framework. Covariates including age, sex, BMI, and the first five genetic principal components (PC1–PC5) were incorporated to control for confounding factors such as population structure. The results of the MiST analysis indicated that Pπ represents the p-value assessing the overall effect of rare *SREK1* variants on obesity risk, while π^ denotes the estimated cumulative effect size of these variants. Pτ assesses the residual heterogeneity among the analyzed variants, indicating whether there is significant variability in their effects beyond the collective association observed. The detailed protocol has been described elsewhere (41)

### Structure Analysis of SREK1

The domain composition SREK1 protein is taken from InterPro database entry Q8WXA9. The structural model of the RRM domains is extracted from AlphaFold database, entry A0A2I2YTW4 (gorilla sequence, the human splice variant with 2 RRM domains is absent in UniProt and consequently also in AlphaFold).

### Cell lines and routine cell culture

Human KOLF2.1J embryonic stem cell lines were maintained in supplemented StemFlex media (Thermo Fisher Scientific A3349401) in freeder-free conditions on a basal matrix of Geltrex (Thermo Fisher Scientific A1413202). TrypLE Express (Gibco, 12604021) and 10μM ROCK Inhibitor Y-27632 dihydrochloride (Stemcell Technologies, 72304) was used for routine maintenance splitting. The absence of mycoplasma was confirmed using an EZ-PCR Mycoplasma Test Kit (Biological Industries, 20-700-20) according to the manufacturer’s instructions.

### CRISPR-Cas9-mediated targeting of SREK1

Three different guide RNA was designed using the CRISPick (https://portals.broadinstitute.org/gppx/crispick/public). Selection of guide RNA was based on the proximity of the cleavage site and the desired gene editing site. Additional criteria include the highest chance of successful targeting and the least chance of off target editing reported by the program. For the production of guide RNAs, a 120 nucleotide oligo (Integrated DNA Technologies Inc.) including the SP6 promoter, guide RNA sequences, and scaffold region were used as a template for synthesis by *in vitro* transcription using the MEGAscript SP6 kit (Thermo Fisher, AM1330) as previously described (21). Since guide RNAs vary in their efficacy, the relative cutting efficiencies of the three guide RNAs were tested in *in vitro* cleavage assays and we selected the guide RNAs that showed activity at the lowest Cas9 concentration for transfection in the Kolf2.1J cells. The HDR templates were designed by using the IDT Alt-R CRISPR HDR Design Tool (https://www.idtdna.com/pages/tools/alt-r-crispr-hdr-design-tool). These single-stranded oligodeoxynucleotides (ssODN) templates are of 100bp containing target variants in the middle and silent variants within PAM sites. The silent variant within the PAM site in the repair template will prevent re-editing and improve editing efficiency.

### CRISPR-Cas9 ribonucleoprotein (RNP) complex-mediated editing in iPSCs

To genetically edit the KOLF2.1J cells by homology-directed repair (HDR), 3 μg purified sgRNA was mixed with 4 μg of recombinant Cas9 nuclease (IDT 1081060) for 45 min at room temperature to form stable RNP complexes. The complex together with 1 μl of 100 μM ssODN was then transferred to a 20μl single-cell suspension of 2 × 10^5^ cells in P3 nucleofection solution and electroporated using Amaxa 4D-Nucleofector™ (Lonza) with program CA137. Transfected cells were seeded onto Geltrex-coated 24 well plates containing a pre-warmed StemFlex medium containing Revitacell (100x, Gibco A2644501) and Penicillin/Streptavidin (ThermoFisher Scientific, 15140-122). HDR enhancer (IDT 1081072) was added to the cells at a 30 μM final concentration for each well. The following day medium was changed to growth medium without Pen/Strep and Revitacell. To increase HDR efficiency, cells were cultured under cold shock conditions (32°C at 5% CO_2_ in air atmosphere) for 48hr post transfection. Cells were given approximately 5-6 days to recover before single cells were then distributed into multiple Geltrex (1:40)-coated 96 well plates by an Aria-Fusion sorter with a 100 μm nozzle. After ∼2 weeks, viable clonally-derived colonies were consolidated into duplicate 96 well plates to allow parallel cell cryopreservation and genomic DNA (gDNA) extraction for multiplex sequencing using Fluidigm index primers followed by MiSeq Nano Technology as previously described (42). Briefly, gDNA was extracted using HotShot buffer and the target regions were amplified using locus-specific primers to generate amplicons approximately 150-200 bp in length. These “first-round” primers contained universal Fluidigm linker sequences at their 5’-end.

In the second round of PCR (indexing PCR), Fluidigm barcoding primers were attached to the amplicons to uniquely identify each clone. For sequencing library preparation, barcoded PCR products were combined in equal proportion based on estimation of band intensity on a 2% agarose gel, and the combined pool of PCR products was purified in a single tube using Ampure XP beads (Invitrogen 123.21D) at 1:1 (V/V) to the pooled sample and eluted in 25μl of water according to the manufacturer’s instructions. Library purity was confirmed by nanodrop, and final library concentration was measured using the Agilent Bioanalyzer (High Sensitivity Kit, Agilent 5067-4626) and diluted to 20 nM. Pooled libraries could be combined with other library pools adjusted to 20 nM, and the resulting “superpool” volume was adjusted to a final volume of 20 μl before sequencing which is performed by the Genomics Core, Cancer Research UK Cambridge Institute. Homozygous and WT clones were identified using the open-access workflow “GeneditID” (https://genedotod/gotjib/op/) (42).

Karyotyping was performed on the identified clones using G-banding by the University of Cambridge Core Laboratory shared services at the Stemnovate Ltd on Babraham Research Campus.

### Hypothalamic neuron differentiation of iPSCs carrying SREK1 variant of interest

We utilised a chemically defined medium for greater reproducibility and improved neuronal maturation, based on previously published protocols (21, 43, 44). Briefly, cells with desired variants were maintained in the absence of antibiotics with feeder-free media StemFlex plus 10 μM ROCK inhibitor (Thermo Fisher Scientific, A3349401) and were cultured overnight on 10cm plate Geltrex coated plates (9.5 x10^5^ cells/well for 6-well plates). Next day, neuroectoderm differentiation was initiated by dual SMAD inhibition using XAV939 (Stemgent 04-1946), LDN 193289 (Stemgent 04-0074) and SB 431542 (Sigma Aldrich S4317) and Wnt signaling inhibition using XAV939 (Stemgent 04-1946) in an in-house neural differentiation N2B27 medium (5). From day 2 to day 7, cells were direct toward ventral diencephalon with Sonic hedgehog activation by the addition of Smoothened agonist SAG (1μM Thermo Fisher Scientific 56-666) and purmorphamine (PMC, 1 μM Thermo Fisher Scientific 54-022) with SMAD and Wnt inhibition molecules gradually replaced with N2 B27medium changed every 2 days. At Day 8, the cells were switched into N2B27 with 5 μM DAPT (Sigma Aldrich D5942) to exit cell cycle. On Day 14, the cells were harvested with TrypLE^TM^ supplemented with papain (Worthington LK003176) and re-plated onto laminin–coated 6-well plates at a density of 3×10^6^ cells per well in the presence of maturation medium containing brain-derived neurotrophic factor BDNF (10ng/ml, Sigma) containing N2B27. On day 16 media was changed to Synaptojuice 1 (N2B27, 10ng/ml BDNF, 2 μM PD0332991 (Sigma Aldrich, PZ0199), 5 μM DAPT, 370 μM CaCl_2_ (Sigma Aldrich, 21115), 1 μM LM22A4 (Tocris, 4607), 2 μM CHIR99021 (Cell Guidance Systems, SM13), 300 μM GABA (Tocris, 0344), 10 μM NKH447 (Sigma Aldrich, N3290)). Cells were maintained in Synaptojuice 1 for a week before changed to Synaptojuice 2 (N2B27, 10 ng/ml BDNF, 2 μM, 370 μM CaCl_2_ 1 μM LM22A4, 2 μM CHIR99021). Cells were then maintained in Synaptojuice 2 until day 36, with media renewal every second day throughout the differentiation and maturation period.

### Fluorescence-activated cell sorting of neurones

Based on the identity of cell surface signature (45), we performed an unbiased fluorescence-activated cell sorting (FACS) on the Kolf2.1J-differentiated hypothalamic neuronal culture to enrich for neuronal populations. By the use of a combination of four cell surface marker antibodies, post mitotic neurons (CD184−/CD44−/CD15LOW/CD24+) were enriched from neuron progenitors and glia cell in the differentiated cultures.

Briefly, day-28 differentiated and matured hypothalamic neurons were dissociated and suspended in the FACS buffer (1%BSA, 2mM EDTA in PBS). Cells were then filtered through a 5-ml polystyrene round-bottom tube with cell-strainer cap (Falcon) and FACS analysed with a BD Biosciences ARIA-II Cell Sort.

### Differential gene expression analysis

RNA was isolated from differentiated hypothalamic cells using the QIAGEN RNAeasy Plus Micro Kit. RNA quality was assessed using the Agilent 2100 Bioanalyzer and sequencing libraries were generated using the Takara Clonetech SMARTer Stranded Total RNA-Seq Kit V2. The libraries were multiplexed and sequenced on an Illumina NovaSeq 6000 instrument with a minimum of 30 million reads per sample. Sequence mapping and gene-level abundance to GRCh38, Ensembl gene model version 100 was determined using STAR 2.5.0a, and differential gene expression was performed with EdgeR’s Quasi-likelihood f-test with a False Discovery Rate of <0.05. All fastq data were deposited in the NCBI’s Gene Expression Omnibus. The experimental design of the gene-editing pipeline is illustrated in Supplemental Figure 2.

### Quantitative Real Time PCR

Expression changes in SNORD115 and SNORD116 were validated through quantitative real-time polymerase chain reaction (qPCR). Specific primers for group I and II of SNORD116 and SNORD115 were designed according to literature (46). All samples were amplified in triplicates and reverse transcribed using the SuperScript™ IV Reverse Transcriptase (Invitrogen, 18090050) following the manufacturer protocol. RNA expression level was quantified by real-time quantitative PCR (RT–qPCR) using the SYBR^TM^ Green universal master mix (Applied Biosystem 4309155) and performed in QuantStudio 5 (Thermo Fisher Scientific.) with an initial hot start of 95°C for 15 min followed by 45 cycles of 95°C for 15 s and 60°C for 30 s. In order to normalise qPCR reactions, GAPDH (Glyceraldehyde 3-phosphate dehydrogenase) was included as housekeeping gene. The relative expression of genes was calculated based on the average Ct values across samples and significance was determined using the multiple unpaired *t*-test.

### Detailed clinical assessment on hyperphagia and PWS

Hyperphagia was assessed using the hyperphagia questionnaire from Dykens et al., which was originally designed and validated for patients with PWS to assess their specific food-related preoccupations and problems (23). The physician completed the questionnaire by asking questions to the parents and/or the proband. The Dykens Hyperphagia Questionnaire includes 13 items, each rated on a 5-point Likert scale where “0” indicates “not a problem” and “5” indicates a “severe/frequent problem.” Three sub-scores are calculated: hyperphagic behavior, food-seeking behavior, and food-related preoccupations.

### Clinical diagnostic criteria for PWS used that devised by Holm et al (25)

There are three categories of diagnostic criteria for PWS: major, minor and supportive, and these are scored on a weighted point system. Major criteria are valued at one point and minor at one half point. Supportive criteria are not included in the point system but serve to increase the confidence in the results determined by the other criteria. In children 3 years of age and younger, only 5 points are required for diagnosis, four of those must come from the major group. A total score of eight is necessary for the diagnosis in the 3 years to adulthood group. In this age group, major criteria items must comprise five or more points of the total score.

### Proband detailed biochemical analysis

Metabolic markers, including plasma/serum leptin and insulin were measured using commercially available ELISA kits from Monobind (Lake Forest, USA). Blood glucose, HbA1c, lipids (cholesterol and triglycerides), markers for thyroid function (free thyroxine, TSH), gonadal function (testosterone, oestradiol, LH, FSH), renal function (electrolytes), and liver function (ALT, AST, gGT) were determined using commercial chemistry analysers.

### Statistics

For statistical analyses, GraphPad Prism 7, Microsoft Excel, and R were used. All data involving a statistical analysis being reported met the criteria to use the appropriate statistical tests. Statistical tests used are reported in the figure legends. Variant calling was performed using the Genome Analysis Toolkit (GATK) following best practices. Variants were filtered based on depth of coverage (>20x) and Phred-scaled quality scores (>30). Annotation was conducted using Ensembl Variant Effect Predictor (VEP), with population allele frequencies obtained from gnomAD and ClinVar for pathogenicity assessment. Differential gene expression analysis, if applicable, was conducted using EdgeR’s Quasi-likelihood F-test with a significance threshold of False Discovery Rate (FDR) < 0.05. Statistical significance for qPCR experiments was determined using multiple unpaired t-tests, with significance thresholds of p<0.01 and p<0.05, as indicated in the figure legends.

### Study approval

The clinical studies were approved by the institutional and hospital ethical committees, and written informed consent was obtained from all study participants or their legal guardians. All procedures involving human subjects adhered to the Declaration of Helsinki.

## Data availability

Data will be made available upon reasonable request to the corresponding authors.

## AUTHORS CONTRIBUTION

SS, AB, PF, YCLT and GSHY conceptualized and designed the study. Methodology was developed by SS, AMS, YCLT, AB, PF, and GSHY. RK, QMJ, JM, HA, WIK, and MA were responsible for recruiting participants, as well as preparing and collecting the data. Experiments and formal data analysis were conducted by SS, AMS, YCLT, MD, BT, BYHL, SA, EV, EBF, SA, and AB. VS performed the protein modelling experiments. NR conducted detailed psychological analysis of the patients. APG contributed by comparing data with PWS patients and provided the necessary forms for this comparison. SOR provided expert clinical insights on the diagnosis and data interpretation. The first draft of the manuscript was written by SS and YCLT. The manuscript was finalized collaboratively by SS, YCLT, SO, AB, PF, and GSHY. All authors reviewed, revised, and approved the final draft.

## Supporting information

Supplemetal Infomation

## ACKNOWLEGDMENTS

We thank Frédéric Allegaert, Timothée Beke and Stefan Gaget (INSERM UMR 1283, CNRS UMR 8199, University of Lille, Lille, France) for technical assistance. We also thanks Mathilde Boissel and Lijiao Ning (INSERM UMR 1283, CNRS UMR 8199, University of Lille, Lille, France) for help with the statistical analyses. The PROMIS data used for analysis described in this manuscript were obtained from the database of Genotypes and Phenotypes (dbGaP) through dbGaP accession number phs000917. The burden analysis was performed using data from the UK Biobank (Application #67575). We extend our gratitude to the patients and their families for their participation in the study.

## Conflict-of-interest statemen

The authors have declared that no conflict of interest exists

## Data Availability

All data produced in the present study are available upon reasonable request to the authors

