## Supplementary material for "Biallelic variants in *SREK1* downregulating *SNORD115* and *SNORD116* cause a novel Prader-Willi-like syndrome": Supplemetal Infomation

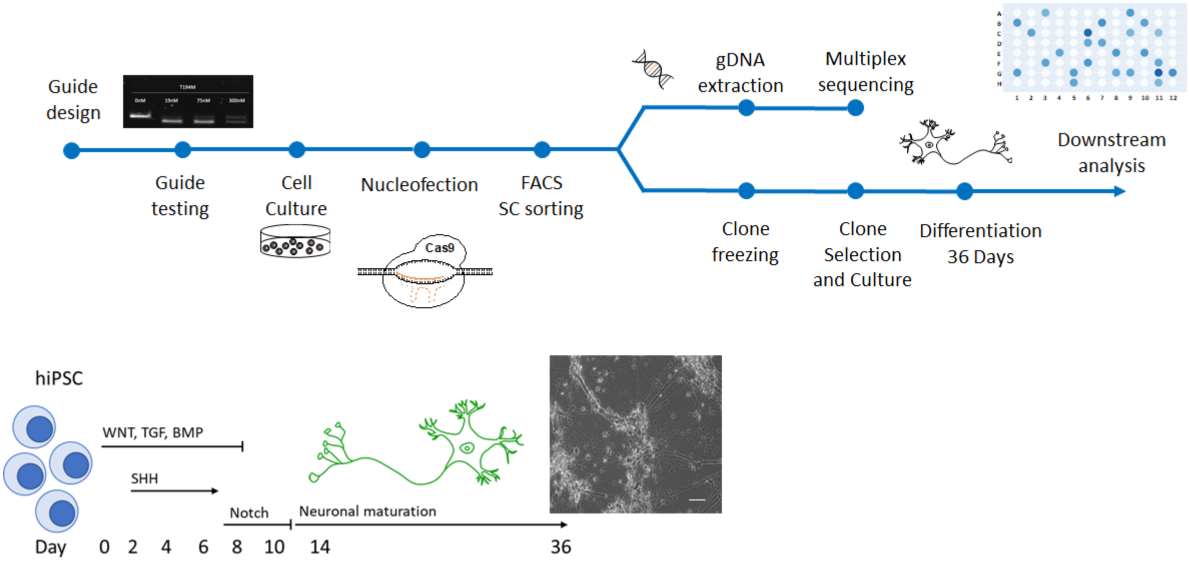


**Figure S1: Experimental design of gene-editing pipeline**

**(A)** The schematic outlines the experimental workflow, starting with *in vitro* guide RNA testing for targeted ssODN-mediated mutation knock-ins. Following CRISPR gene targeting, iPSC clones were isolated using FACS into 96-well plates, which were duplicated to support clone propagation and the barcoded PCR amplification of the *SREK1* locus to facilitate deep sequencing to identify clones harbouring the mutations of interest. **(B)** Selected clones, identified using GenEditID, were expanded and cryopreserved before undergoing a 36-day neuronal differentiation protocol. Downstream analysis was performed after FACS, which enriched neuronal populations using a combination of four cell surface marker antibodies.

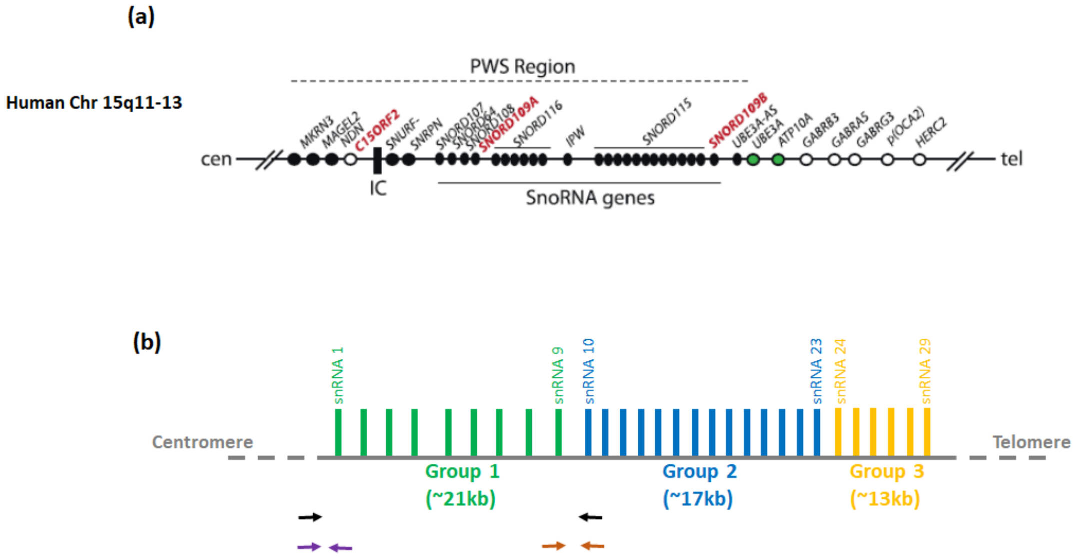


**Figure S2: Prader-Willi syndrome (PWS) locus**

**(A)** In human, the critical region for PWS is located on paternal chromosome 15 and contains several known protein coding genes (e.g. *MAGEL2*, *SNURF-SNRPN*). Six snoRNAs have been reported within this locus, which include 29 copies of *SNORD116* (29 copies) and 48 copies of *SNORD115*. **(B)** Within *SNORD116* loci, the 29 copies of human *SNORD116* can be divided into 3 distinct groups by sequence homology, with group I expression significantly higher than group II or III in the hypothalamus.

**Supplementary Table 1:** Null variations detected in the SREK1 gene (NP_001070667.1) among 187,242 participants from the UK Biobank, analyzed in the study.

| Variations | Consequence | Variant class |
|---|---|---|
| p.Arg307Ter | stop_gained | SNV |
| p.Arg548Ter | stop_gained | SNV |
| p.Glu549ThrfsTer10 | frameshift_variant | deletion |
| p.Arg463ProfsTer4 | frameshift_variant | deletion |
| p.Arg548AsnfsTer6 | frameshift_variant | deletion |
